# is metallic perception a taste, an aroma or a flavor ?

**DOI:** 10.1101/2024.09.13.24313668

**Authors:** Guillaume Buiret, Thomas-Danguin Thierry, Feron Gilles

## Abstract

**Aim:** Metallic taste is known to vanish with nasal occlusion, suggesting it should be termed metallic “aroma” rather than metallic “taste.” However, it also appears after neurological injuries, such as a chorda tympani section, which suppresses taste perception mediated by the facial nerve.

**Methods:** In 120 healthy volunteers, the perception of an iron sulfate solution was assessed at different lingual locations—corresponding to the facial and glossopharyngeal nerves—and with either open (n=60) or closed (n=60) noses to evaluate if it is a taste or an aroma.

**Results:** Nasal occlusion significantly reduced the perception of iron sulfate. It prevented perception in 31.7% of participants, though it did not completely eliminate it in the remaining 68.3%. Additionally, in open-nosed subjects, the intensity of iron sulfate perception was significantly greater when applied to the base of the tongue (glossopharyngeal nerve) than to the tip (facial nerve). These differences did not persist with nasal occlusion.

**Conclusion:** Nasal occlusion diminished but did not abolish the perception of iron sulfate. With an open nose, a differential taste perception was significant, but not with a closed nose. Therefore, metallic perception involves both retro-olfactory and gustatory components, suggesting it is a metallic flavor.

## INTRODUCTION

The prevalence of metallic taste (MT) affects approximately 29% of people undergoing cancer treatment (1). MT is known to vanish with nasal occlusion, leading some authors to suggest a retro-olfaction mechanism (2-7). However, its perception also depends on neurological factors. Taste is mediated by two nerves: the facial nerve for the anterior tongue and the glossopharyngeal nerve for the posterior tongue. The release of facial nerve inhibition on the glossopharyngeal nerve (8-14) could be a potential cause of MT. This hypothesis has been proposed to explain the appearance of metallic taste after chorda tympani sectioning—a branch of the facial nerve—during middle ear surgery (11-14).

Under normal circumstances, the facial nerve appears to exert an inhibitory effect on the glossopharyngeal nerve, thus limiting taste perception in the posterior part of the tongue. When the chorda tympani is severed, the abrupt cessation of sensory input to the anterior tongue causes this inhibition to be lifted, possibly explaining MT perception. By analogy, anticancer treatments, which can cause neurotoxicity (15), might also release facial nerve inhibition on the glossopharyngeal nerve, leading to MT in these circumstances.

While much data is available on the overall perception of iron sulfate (2-6, 16-21), insufficient data exists on tests at precise locations on the tongue to distinguish perceptions mediated by the facial or glossopharyngeal nerves. So, is metallic “taste” a taste or an aroma?

We questioned whether differences in the perception of induced metallic taste exist depending on the test’s location in the oral cavity and on retro-olfaction. To answer this, we assessed the difference in the perception of an iron sulfate solution at different lingual locations (facial and glossopharyngeal nerve territories) and whether the nose was open/occluded in healthy volunteers.

## METHODS

### Objective

The primary objective of this study was to evaluate the variability in the intensity of iron sulfate perception based on tongue location in healthy volunteers with and without nasal occlusion.

### Recruitment

Patients or accompanying adults in the waiting room of the ENT consultation department were offered the opportunity to participate in supraliminal tests of iron sulfate perception intensity. Voluntary adults were included, while non-inclusion criteria were a history of head and neck cancer, consultations for taste/smell disorders, refusal to participate, minors, adults under guardianship, or insufficient French language proficiency to answer the questions. No exclusion criteria were applied.

Tests were conducted without nasal occlusion in the first 60 participants (50%) and with nasal occlusion in the last 60 (50%) to prevent retro-olfactory airflow. The study complies with the Declaration of Helsinki for Medical Research involving Human Subjects and was authorized by the CPP Est IV on 16/12/2021 (no. 2021-A02149-32) for the open nose and on 05/08/2022 (no. 2022-A01475-38) for the occluded nose. All participants provided informed written or verbal consent. The study has been registered on Clinicaltrials.gov (NCT05227157).

### Data Collection

Data collected included gender, age, smoking status, open/occluded nose status, and quantification of supraliminal sensory intensity testing.

### Testing Process

After confirming eligibility and written consent, a cotton swab soaked in a 10 g/L iron sulfate solution diluted in Evian water was applied to each predefined lingual location (see Figure 1 and video). The order of tests were always from A to D. A procedure was performed to dilute 250mg of iron sulfate in 25ml of mineral water. The supplier of the iron sulfate was Supelco (ref #1.03965). The glossopharyngeal nerve mediates taste at the posterior tongue (base), and the facial nerve mediates taste at the anterior tongue (tip). Participants rated the perceived intensity on a scale from 0 (no perception) to 5 (highest imaginable intensity) at each of the four predefined lingual locations. The mouth was rinsed with Evian water between tests; patients had to split out the mouthrinse, not to swallow it.

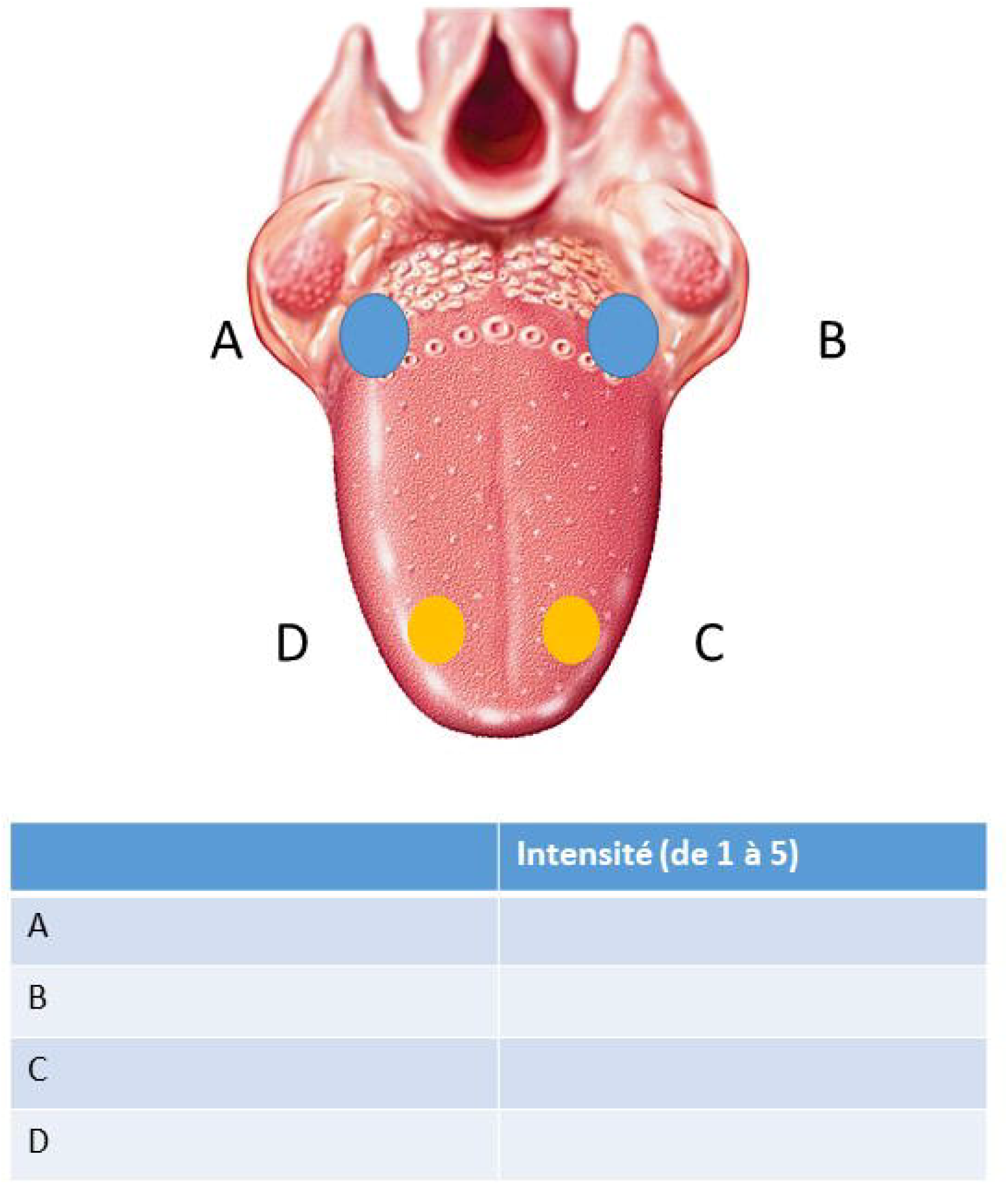

### Data Analysis

- Group comparability was ensured by comparing age means using Student’s t-test and qualitative data using the chi-square test.
- In the overall population, and sub-populations with open/occluded noses, arithmetic means were compared for quantitative data from supraliminal sensory tests: i. between each site (left front, right front, left back, right back), ii. between the right and left sides, and iii. between the anterior and posterior parts of the tongue.
- Means for each of the four sites were then compared according to open or occluded nose status.

## RESULTS

A total of 120 participants were recruited from December 16, 2021, to August 8, 2023.

In the open nose group, 29 were females (48.3%) and 31 were males (51.7%). In the occluded nose group, 36 were females (60%) and 24 were males (40%). There was no significant difference in gender distribution between the two groups (p=0.2717). Figure 2 presents the age distribution according to nose occlusion status. Participants in the open nose group were significantly older than those in the occluded nose group (mean age 47.9 ± 18.8 years and 39.0 ± 16.0 years, respectively; p=0.0056). Tobacco consumption between the two groups was not significantly different (p=0.06559).

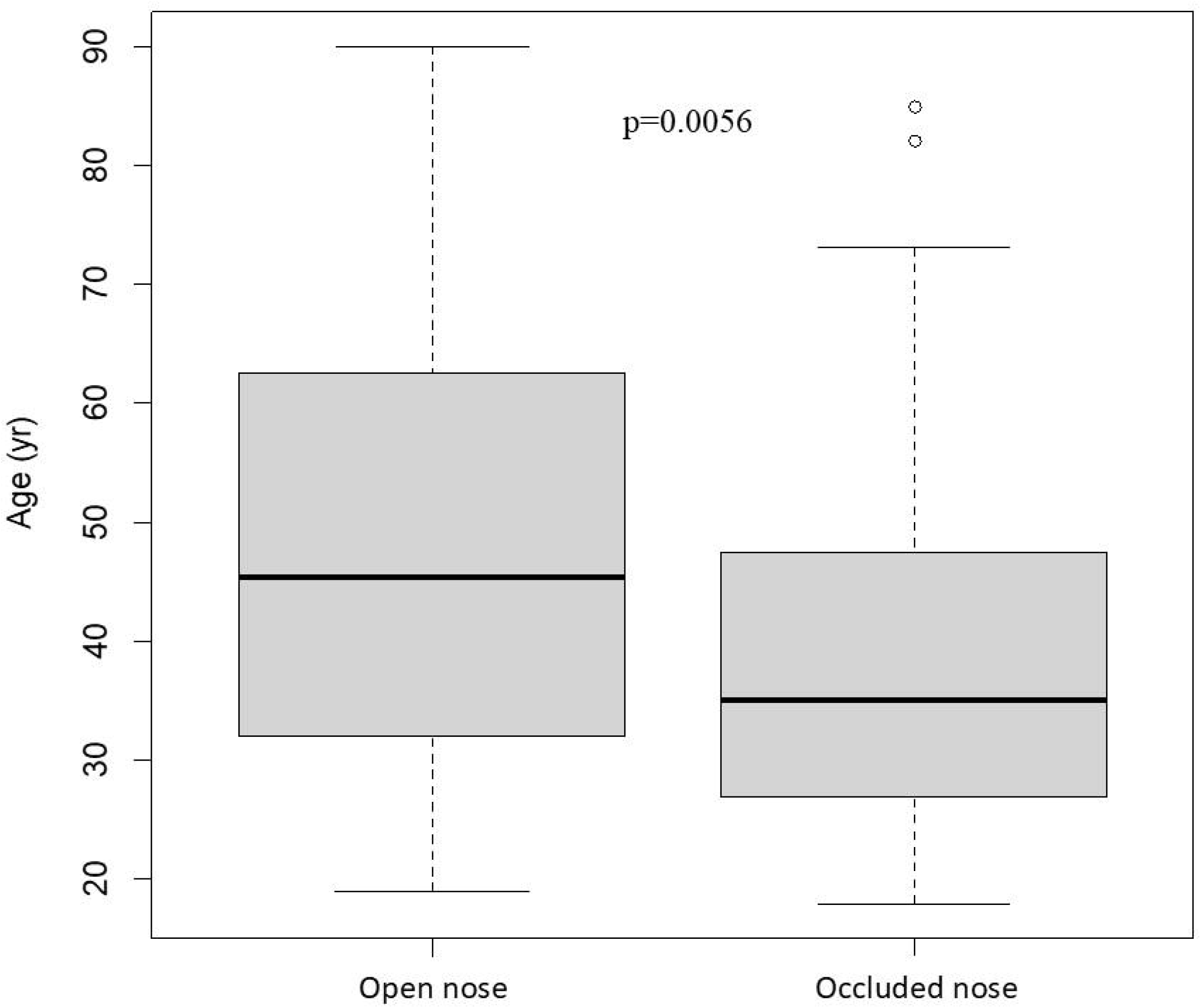

Figure 3 presents the intensity test results of MT perception in the subpopulation without nasal occlusion (n=60) and with nasal occlusion (n=60), according to test location. Comparison by location according to open/occluded nose status showed that all perceptions at each location were significantly higher when the nose was open (p<10^−5^ for right back, left back, left front, and right front).

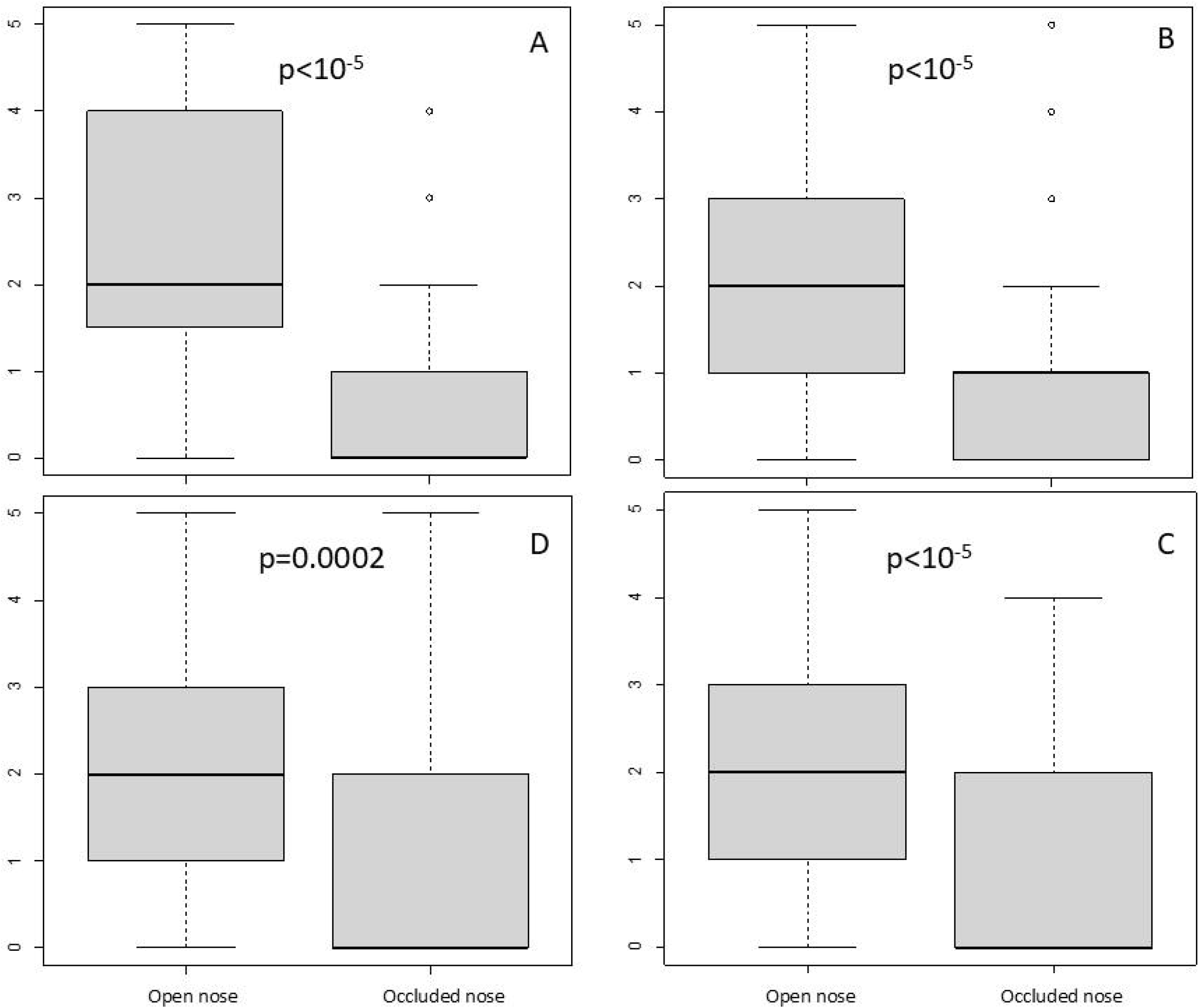

Figure 4 presents the intensity test results of MT perception in the subpopulation without nasal occlusion (n=60) and with nasal occlusion (n=60), according to the sum of test locations: left, right, front, and back. All perceptions at each location were significantly higher with an open nose (p<10^−5^). Among the 60 participants without nasal occlusion, only one had no perception of the iron sulfate solution in any location (1.7%). Comparison of perceptions on the right and left sides showed a non-significant difference in means of -0.4/5 (p=0.0603). Comparison between perceptions at the front and back of the tongue showed a significant difference in means of -0.76/5 (p=0.02014): intensity was significantly higher at the base of the tongue than at the tip.

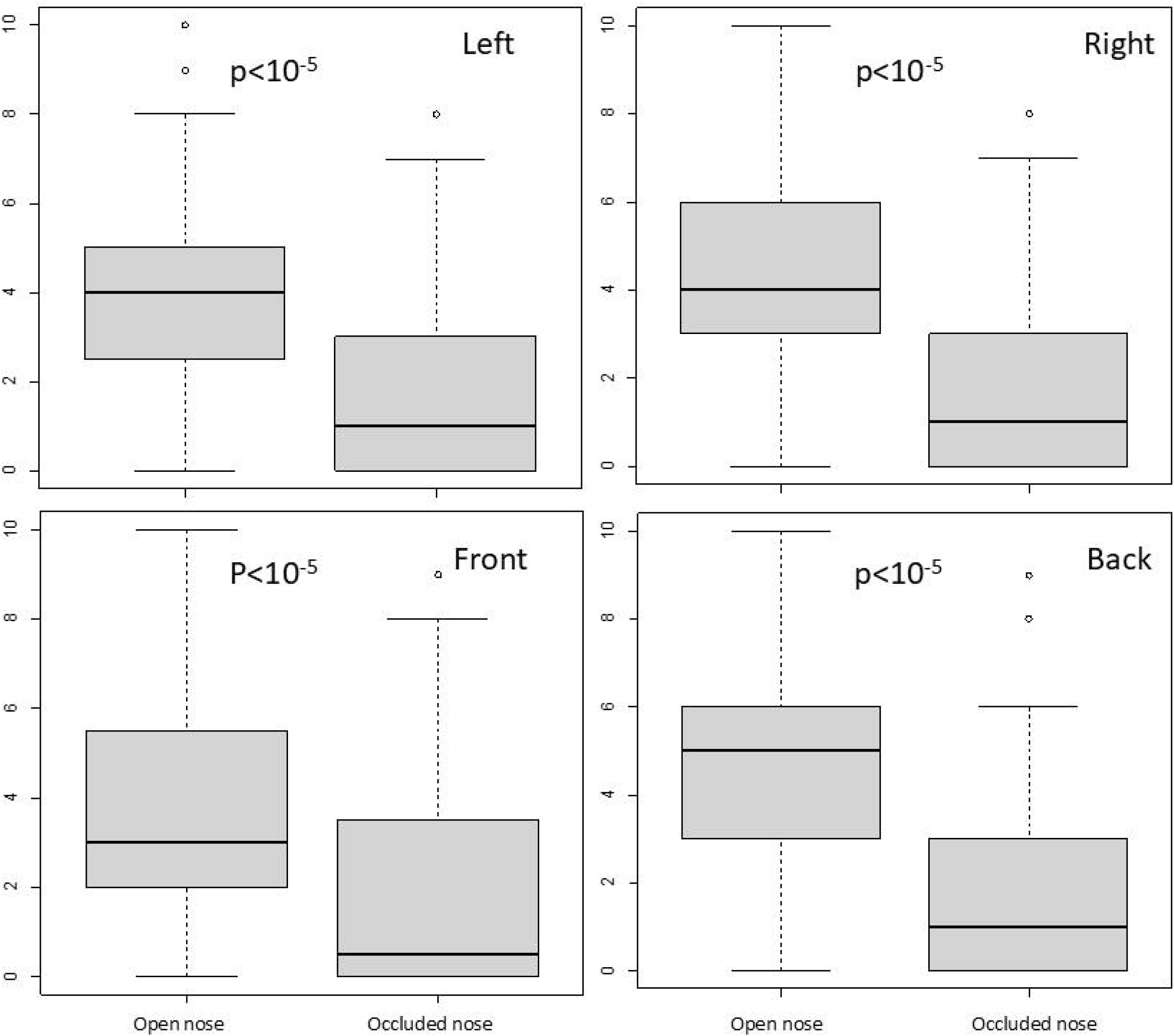

Among the 60 participants with nasal occlusion, 19 had no perception of the iron sulfate solution in any location (31.7%). Comparison of perceptions on the right and left sides showed a non-significant difference in means of -0.18/5 (p=0.5704). Comparison between perceptions at the front and back of the tongue showed a non-significant difference in means of +0.4/5 (p=0.1914): intensity was higher at the tip of the tongue than at the base.

## DISCUSSION

We were able to show, in healthy volunteers: i. that nasal occlusion significantly reduced the perception of iron sulfate (it even prevented it in 24% of people, without causing it to disappear in the others); ii. that the intensity of perception of iron sulfate was significantly greater when the product was applied to the base of the tongue than to the tip, whereas there was no significant difference between the right and left sides in open-nose subjects; iii. that the latter differences did not persist in the case of nasal occlusion. These results are consistent with those published by other teams, who have also shown that nasal occlusion increases iron sulfate detection thresholds (2, 3, 5, 6, 16, 19, 22) without making it disappear. Only Hettinger et al. (4), in 12 healthy subjects, found no perception of iron sulfate during nasal occlusion. We can deduce that the perception of iron sulfate is mainly linked to olfaction, since nasal occlusion drastically reduces its perception. This is concordant with the report of Lim and Lawless explored the perception of ten divalent salts. They showed that the iron sulfate was plotted away from the four primary tastes domain when the nose was open and this difference disappeared when the nose was closed. This difference was therefore caused by the smell (22). Smell can also explain the differences in perception between our groups, regardless of the territory, between the anterior and posterior parts of the tongue. With the nose open, perception at the base of the tongue (closer to the back of the nose, i.e., perception by retro-olfaction) should counterbalance the intensity differential between the front and back of the tongue. However, this perception was not completely abolished in the majority of patients (76%) during nasal occlusion, despite the 0.5/10 mean difference in gustatory perception between the territory of the glossopharyngeal nerve (base of the tongue) and the territory of the facial nerve (tip of the tongue) no longer being significant (p=0.08021).

There may be two explanations: the perception of iron sulfate may be a flavor in its own right, essentially combining mainly retro-olfactory perception with other components of the perception (taste, chemosensory). Indeed, Riera et al showed a direct response of TRPV1 receptor to metallic salts in vitro (23). Omur-Ozbek et al. also suggested a potential role for trigeminal stimulation. In their study, Fe^2+^ was perceived as astringent during nasal occlusion and then as merely metallic after removal of the nasal clips (19). Similarly, Stevens et al., with their concept of multidimensional taste scales applied to iron sulfate in 47 healthy volunteers, showed that metallic taste was not grouped with traditional sapid flavors (the closest flavor was bitter) but rather with oral chemical sensations such as aluminum (astringency, trigeminal sensitivity) and glutamate (umami flavor), even after nasal occlusion (21).

Another explanation could be linked to the metal’s olfactory detection threshold, such as 1-octen-3-one and 1-nonen-3-one (22), trans-4,5-epoxydecenal, (Z)-1,5-octadien-3-one and 1-octen-3-one (24-27), and epoxydecenal (28), which could be acute, on the order of parts per billion. In this case, incomplete nasal occlusion should have occurred in the 74% of people with nasal occlusion who still perceive metallic taste. But in that case, as the tongue base is closer to the retronasal pathway than the tip of the tongue, the intensity should be higher at the tongue base than at the tip of the tongue, which was not highlighted in our data (Figures 3 and 4).

Finally, other research supports the notion that MT perception involves oral somesthesia. Studies by Stevens and colleagues using multidimensional taste scales have shown that metallic taste was associated with oral chemical sensations such as astringency and trigeminal sensitivity, rather than traditional gustatory qualities like sweet, sour, salty, and bitter (21). Additionally, Hettinger et al. demonstrated the role of olfaction and somatosensory pathways in the perception of non-traditional taste stimuli, further highlighting the complexity of metallic taste perception (4). These findings suggest that metallic taste may be a multidimensional sensory experience involving both olfactory and trigeminal contributions.

Finally, it is important to clarify certain methodological choices that were made, not as limitations, but as deliberate considerations aimed at ensuring the robustness of our results and minimizing potential biases. Specifically, we chose the use of cotton swab instead of drops to avoid the diffusion of the iron sulfate solution in the whole mouth, especially in the two territories of taste perception.

We also made the choice to conduct the two experiments in two different populations that after the end of the studies were not comparable. Conducting the test with both the nose open and closed on the same individuals could lead to cross-contamination of taste perception. When the solution is applied once in a given territory, the lingering taste or aftereffects might influence the subsequent measurements with the nose closed. This would compromise the clarity and independence of the results. By using independent groups, we wanted to avoid the risk of biased taste responses due to repeated exposure. Repeated exposure to the same stimulus could result in taste adaptation, where the participants’ sensitivity to the solution diminishes over time. By splitting the participants into two independent groups, the risk of this kind of sensory fatigue was minimized, maintaining the integrity of the data. Despite it has been reported that metallic sensation following oral stimulation of ferrous sulfate is most likely due to retronasal smell, following lipid oxidation in the mouth (6). End products of lipid oxidation are typically hard to remove by water rinsing itself and often leave lingering sensation. However Evian water is the most neutral mineral water and is widely used to dilute testants.

## Conclusion

This study demonstrated that nasal occlusion significantly reduced the perception of iron sulfate, confirming that this perception was primarily linked to olfaction. Perception varied depending on the tongue’s position, with higher intensity at the base compared to the tip, a difference nullified by nasal occlusion. Despite occlusion, 76% of subjects still perceived the metallic taste, suggesting a possible involvement of a real metallic taste, trigeminal stimulation, or very low olfactory detection thresholds.

## Data Availability

All data produced in the present study are available upon reasonable request to the authors

## Funding to report to this submission

This wok was supported by the Comités de la Drôme et de l’Ardèche de lutte contre le Cancer.

## Aknowledgements

to C.T. Molta, MD, for the English translation and editorial input.

Data is available on request

